# COVID-19 disruptions of food systems and nutrition services in Ethiopia: Evidence of the impacts and policy responses

**DOI:** 10.1101/2022.08.31.22279432

**Authors:** Juliet McCann, Lea Sinno, Eki Ramadhan, Nega Assefa, Hanna Y. Berhane, Isabel Madzorera, Wafaie Fawzi

## Abstract

**Background:** Since its first case of COVID-19 on March 13, 2020 and Ethiopia has exerted efforts to curb the spread of the Coronavirus disease 2019 (COVID-19) without imposing a nationwide lockdown. Globally, COVID-19 related disruptions and mitigation measures have impacted livelihoods and food systems, nutrition, as well as access and use of health services.

**Objective:** To develop a comprehensive understanding of the impacts of the COVID-19 pandemic on food security and maternal and child nutrition and health services and to synthesize lessons from policy responses to the COVID-19 pandemic in Ethiopia.

**Methods:** We conducted a review of literature and 8 key informant interviews across government agencies, donors, and non-governmental organizations (NGOs), to map the impacts of the COVID-19 pandemic on the food and health systems in Ethiopia. We summarized policy responses and identified recommendations for future actions related to the COVID-19 pandemic and other future emergencies.

**Results:** The impacts of the COVID-19 pandemic were felt across the food system. Disruptions were noted in inputs supply due to travel restrictions and closed borders restricting trade, reduced in-person support by agriculture extension workers, income losses, increases in food prices, and the reduction in food security and consumption of less diverse diets. Maternal and child health services were disrupted due to fear of contacting COVID-19, diversion of resources, and lack of personal protective equipment. Disruptions eased over time due to the expansion of social protection, through the Productive Safety Net Program, and the increased outreach and home service provision by the health extension workers.

**Conclusion:** Ethiopia experienced disruptions to food systems and expanded existing social protection and public health infrastructure and leveraged partnerships with non-state actors. Nevertheless, vulnerabilities and gaps remain and there is a need for a long-term strategy that considers the cyclical nature of COVID-19 cases.

## Introduction

Coronavirus disease (COVID-19), caused by infection with SARS-CoV-2, has led to a global public health crisis. The earliest cases were detected in December 2019, and on March 11^th^, 2020, the World Health Organization (WHO) characterized COVID-19 as a pandemic.^1^ As of May 6, 2022, Ethiopia has experienced 470,609 cases and 7,510 deaths due to COVID-19 and 29,373,478 vaccines have been administered.^2^ Before the start of the COVID-19 pandemic, Ethiopia was burdened with poor maternal and child health indicators. Child malnutrition including high rates of stunting (38.4%) and wasting (9.9%) among children under 5 years, and anemia among women of reproductive age (23.6%) are severe public health challenges.^3^ Additionally, in 2019 51.5% of households reported experiencing food insecurity, with the average household not able to satisfy food needs for 1.3 months.^4^ This is especially concerning during this pandemic, since malnourished children are at higher risk of COVID-19 related complications and deaths due to their weakened immune system and households are likely to sink further into food insecurity.^5^

Ethiopia began implementing measures to address the COVID-19 pandemic in January 2020, ahead of most countries and before it detected its first case, by introducing strict passenger-screening protocols at airports and conducting house-to-house screenings for 11 million households.^6^ The government subsequently scaled up its response after the first confirmed case was reported on March 13, 2020, declaring a federal state of emergency from April to September 2020 (Appendix 1). Ethiopia did not implement a nationwide lockdown as other countries, however it relied on less stringent, public health preventative measures.^7^ The country closed international land borders (except with Djibouti to ensure smooth transport of commercial goods), banned inter-regional public transport and public gatherings, closed schools and entertainment venues, and announced social distancing measures.

Globally COVID-19 disruptions on the food systems have varied by context and based on the extent of government lockdowns, and have included impacts on the availability of inputs for agriculture and food supply chains, food prices, and consumers’ income.^11^ East Africa, including Ethiopia, was affected triple burden of simultaneous disasters in 2020, that is COVID-19, flooding, and desert locust swarms, which adversely impacted food and nutrition security.^12^ These factors have the potential to further adversely impact health and nutrition in Ethiopia.

Health systems have also been strained due to the COVID-19 pandemic, which may have profound impacts on maternal and child health.^13^ Ethiopia has lower than recommended health professional to population ratio (0.96 per 1000 population instead of 4.45 per 100 populations) and this raises the issue of an inadequate provision of the basic and regular health services, especially during the pandemic.^14^ Additionally in Sub-Saharan Africa, there were concerns that lockdown measures taken by governments, including travel restrictions limited the availability of transport for populations, restricting ability to access medical services^15^. Staff re-allocation to deal with the COVID-19 pandemic may have also led to limited access to routine health services. Consequently, foregone care will translate into an increase in morbidity and mortality among vulnerable groups such as women and children.^16^ However, research on the reality of these impacts in Ethiopia has been limited. Robust data and research are required to determine policy actions and review the impact of previous policies and how they can be refined in this context.

We conducted a descriptive case study to develop a comprehensive understanding of the impacts of the COVID-19 pandemic on food security, nutrition, and health in Ethiopia, and assess programs and policies responses to mitigate these impacts. Our objectives were to: 1) examine how the COVID-19 pandemic has influenced the food system in Ethiopia including its effects on agricultural production, food prices, and consumer behaviors; 2) assess the impacts of COVID-19 on health systems and access to maternal, newborn, and child health and nutrition services.

## Methods

### Study setting

Ethiopia is the second-most populous nation on the continent with a population of 115 million people^17^and has the fastest growing economy in the region. Yet, it is also one of the poorest countries.^18^ Approximately 78% of the population resides in rural areas, and of the urban population, 11.3% live in the capital, Addis Ababa. The median age of the population in Ethiopia is 19.5 years.^19^

### Data collection

We conducted a qualitative study in Addis Ababa, Ethiopia between November 2020 and July 2021. We collected data through (a) an extensive literature review including a review of published and grey literature, and b) key informant interviews with stakeholders in Ethiopia.

The literature review included a review of existing literature on food and health systems, as well as on the impact of the COVID-19 pandemic on the food and health systems both in Ethiopia and globally. Due to the rapid development of the pandemic and its impacts, the literature on this intersection between the pandemic and the food and health systems was still emerging at the time of research. Therefore, the research team reviewed the grey literature, newspaper articles, including webinars, project reports, and op-eds written by policy stakeholders. The research team identified work to be reviewed through an independent search and based on recommendations from interviewees.

The research team identified potential interviewees from a list of authors of recent reports on the COVID-19 in Ethiopia, and through contacts in research institutions, universities, and local non-governmental organizations. We recruited informants through snowball sampling by which contacts referred us to experts with knowledge on this topic. Due to travel restrictions, the research team conducted virtual expert interviews with 8 stakeholders based in Ethiopia and the United States. Interviews were conducted from January to July 2021. The full list of interviewees may be found in the Acknowledgments section.

A semi-structured questionnaire was used to conduct interviews. Interviews were conducted by 3 members of the research team (JM, LS and ER) (Appendix 2). During each interview, the research team asked interviewees a series of questions about the impacts of COVID-19 on the food and health systems and the policies implemented to address these disruptions. At each interview, interviewers took notes which were consolidated following the conclusion of the interview.

### Ethical consideration

Ethical approval for the study was sought from the Institutional Review Board (IRB) at the Harvard T.H. Chan School of Public Health, which determined that this work is not human subjects research as defined by the regulations of the U.S. Department of Health and Human Services (DHHS) and Food and Drug Administration (FDA).

## Results

The data collected from the interviews and literature review were summarized. We extracted three common themes from the findings: (a) impacts of the COVID-19 pandemic on the food system (Table 1), (b) impacts on the health system and specifically on maternal and child services (Table 2), and (c) policy responses to the pandemic-related disruptions (Table 3).

**Table 1:** Summary of impacts of COVID-19 on the food systems.

| Topic Area | Impacts |
| --- | --- |
| Agricultural production | • Collaborative agriculture activities such as group work and support from Agricultural Extension Workers were most impacted by the COVID-19 pandemic in Spring 2020 because of concerns about social distancing and a desire to limit interaction.<sup>30,21</sup><br>• One survey in rural Oromia found that almost half of households reported decrease in crop production because of the COVID-19 pandemic.<sup>22</sup><br>• Key informants noted challenges to accessing agriculture inputs such as fertilizer due to price increases; however, more research is needed to determine if these challenges were a direct result of the pandemic or other policy changes. |
| Household income | • Disruptions to business operations and employment due to COVID-19 restrictions caused a decrease in reported income for households across Ethiopia, particularly from May to July 2020.<sup>25</sup><br>• Income losses were most pronounced in urban areas and non-farm businesses, as businesses were not able to operate at full capacity due to risk of COVID-19 transmission.<sup>28</sup> |
| Impacts on food availability, food security, and dietary consumption | • Surveys reported increases in food prices for some staples and crops. Price increase in crops that relied upon import from other countries, such as onions, may be a result of border closures early in the pandemic.<sup>25</sup><br>• The Ethiopian dairy sector has been resilient throughout the pandemic, which may be because this sector is less reliant on input resources than other countries<br>• Food security and dietary diversity was negatively impacted across Ethiopia; however, the Productive Safety Net Program substantially mitigated these impacts for recipient households. Impacts on food security and dietary diversity dissipated over time. |

**Table 2.**
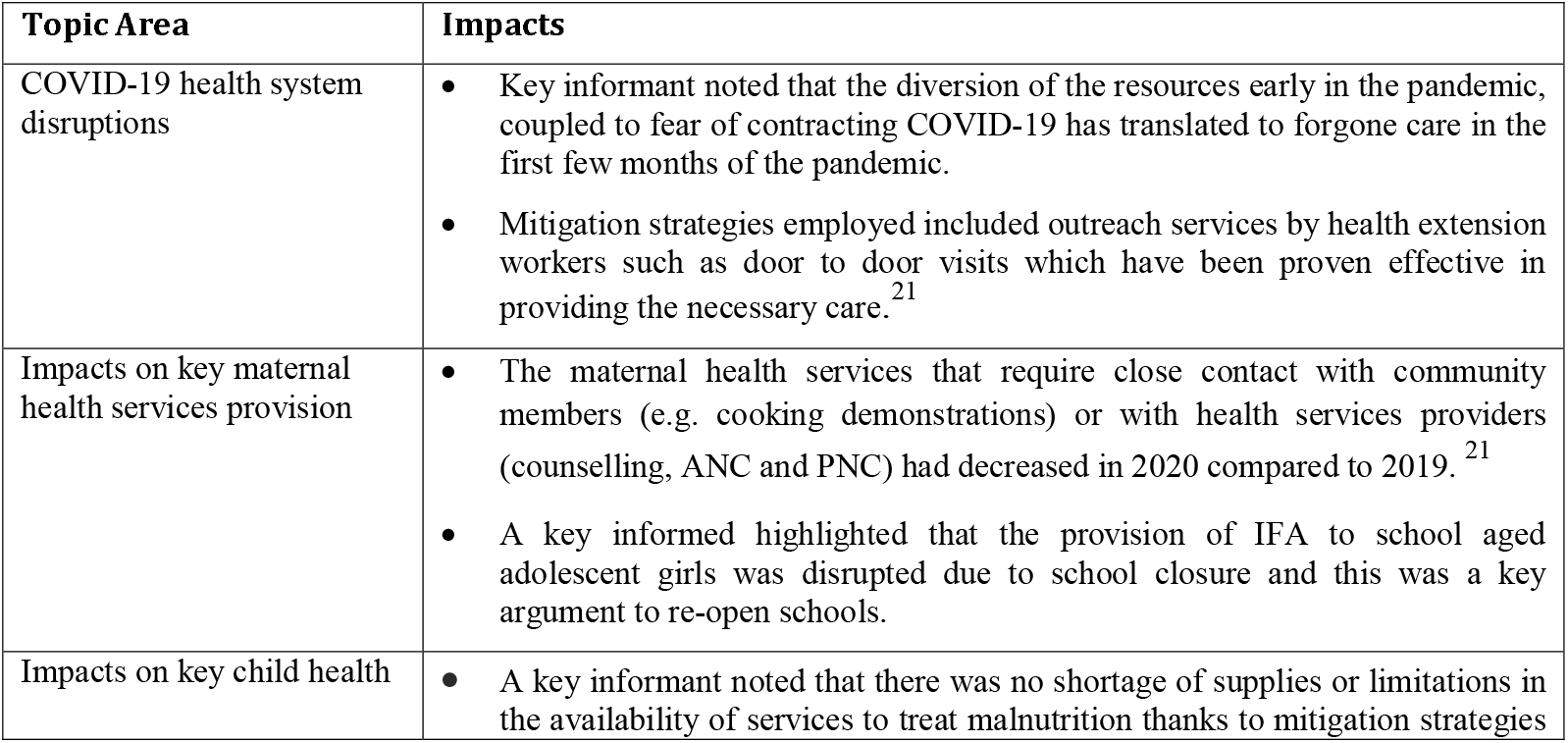
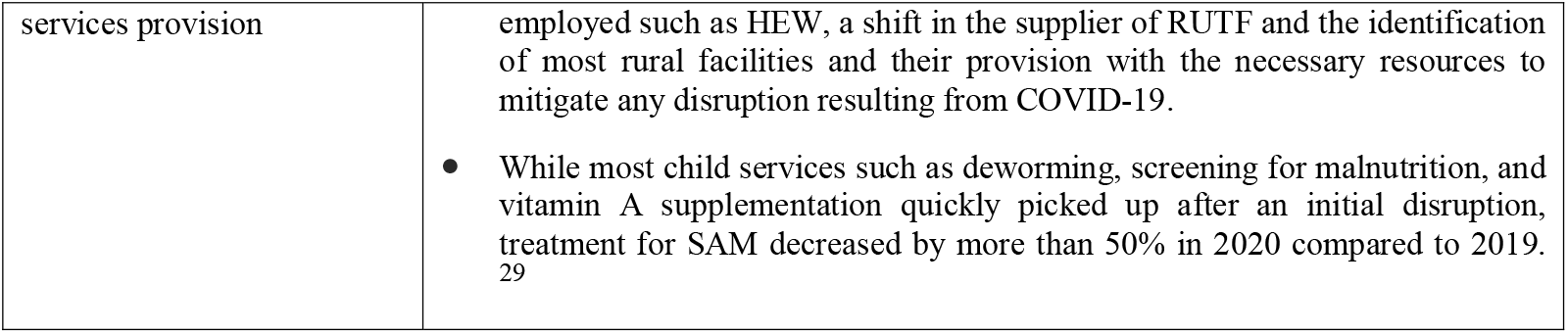
Summary of impacts of COVID-19 on the health system services.

**Table 3.** Summary of policy responses to the impacts of the COVID-19 pandemic on the food and health systems in Ethiopia.

| Program or policy | Implementation Date and Location | Description |
| --- | --- | --- |
| **Health systems** |  |  |
| Training for health extension workers through mobile app | Began in May 2020; nationwide | Ministry of Health collaborated with United States Agency for International Development and Amref Health Africa to deliver immediate training digitally |
| Public health information campaign | July 2020; nationwide | Government and partners disseminated information on the impact of COVID-19 |
| Community Based Action and testing (ComBAT) | August 2020; nationwide | Campaign to curb the spread of COVID-19 by focusing on community-based approaches and mass testing |
| **Food systems** |  |  |
| Cash transfers | April-June 2020; urban areas | Beneficiaries of Urban PSNP received advance 3 months payment |
| Public works expansion | July 2020; rural areas | Rural PSNP scaled up to 1 million people for 3 to 6 months. The cash benefits increased by about 22% |
| Food price controls | March 2020; nationwide | Ministry of Trade and Industry monitored and enforced actions against price gouging by businesses |
| Emergency food distribution | Rural and urban | \$635 million to 15 million individuals vulnerable to food insecurity and not currently covered by the rural and urban PSNP |
| Credit support for farmers | April 2020: nationwide | Development Bank of Ethiopia provided support for micro and small enterprises, microfinance institutions, and cooperatives to provide loans to farmers, businesses, and consumer cooperatives |
| Each One Feed One National Challenge | April 2020: nationwide | Government encouragement for Ethiopians to provide a meal to the most vulnerable sections of society |
| Importation of food free of tax | April 2020: federal level | The government collected bids from international companies to import 17,300 kg rice, 32,000 kg sugar, 104.3 million liters of edible oil |
| Purchase of stockpile of food and essential goods | April 2020: Addis Ababa | Addis Ababa City Administration allocated 600 million Ethiopian birr (ETB) for purchasing of food/other essential goods and distribution to 800 retail shops |
| Food transfers | April 2020: Bahir Dar (Amhara Regional State) | Provision of flour, oil, and sugar to the poorest of the poor |
| Food transfers | April 2020: Adama (Oromia) | Provision of bread and water for those who need assistance during the stay at home order |
| Food rationing program | May 2020: 1000 vulnerable individuals Lideta and Addis Ketema Sub Cities of Addis Ababa | Food distribution program carried out by the Addis Ababa City Administration for three months (supported by the Gates Foundation) |
| Urban agriculture | August 2020: Addis Ababa | Addis Ababa City Administration advocated more urban agriculture to mitigate the impact of food shortages |

### Impacts of the COVID-19 pandemic on food systems in Ethiopia

#### Impacts on agricultural production

The key informant interviews and the literature review outlined possible areas of impact for COVID-19 on collaborative agriculture activities and access to agricultural imports. Key informants highlighted that the impact of COVID –19 on agriculture production, would be felt mostly in rural areas, as agriculture is the main form of livelihood. However, at the time of the interviews in Jan 2021, respondents noted that the rural areas in Ethiopia had few cases of Covid-19 and related disruptions were not likely to be extensive. Nevertheless, one respondent stated the collaborative group work of agriculture has the potential to spread the virus if social distancing is not enforced during such activities. By July 2021, another key informant indicated that farmers had concerns about access to agriculture inputs such as fertilizer because of international price increases. However, the informant noted that the impacts of COVID-19 on these price changes cannot be disentangled from other policies such as changing exchange rates.

Studies examining the impacts of COVID-19 on agriculture activities in Ethiopia reported similar impacts. One study found that within the agricultural sector, field-level agricultural activities that supported social distancing were least affected, however, support from Agricultural Extension Workers and access to input activities were most affected.^21^ The most common reported causes of these disruptions were fear of infection, shortage of inputs, and travel restrictions.^21^ Strategies such as provision of inputs by different institutions and use of SMS messaging were implemented to mitigate the impacts.^21^ In a survey conducted in rural Kersa in Oromia region between July and August 44.8% of households reported that their crop production had decreased because of the COVID-19 pandemic.^22^ However, results from another telephone survey found that farmers in select woredas in the Amhara and Oromia regions reported no major problems in accessing agricultural inputs during the Belg planting season from March to June of 2020.^23^

#### Impacts on food availability and food prices

Although there was no full nationwide lockdown, mobility in parts of Ethiopia was restricted due to disruptions to public transportation. One of the key informants reported that while there was tension among the public, there was not much impact on the food distribution system because there were no restrictions in food transportation. Another key informant similarly observed that food availability for consumer purchase was largely unimpacted after the onset of the COVID-19 pandemic. Key informants highlighted that the impacts of COVID-19 on food security and consumption were not as profound as initially anticipated at the start of the pandemic.

While some surveys reported increases in food prices, the literature review supported the key informants’ reports of minimal changes to dietary consumption because of the COVID-19 pandemic. Phone surveys carried out in Addis Ababa and Kersa documented reported changes in food prices during the COVID-19 pandemic. A cross-sectional phone survey in Kersa with 297 respondents from July to August found that over 95% of households reported that food prices increased from the start of COVID-19 for staples (maize, rice, cassava, and teff), pulses (beans, lentils, peas, and chickpeas), fruits, vegetables and animal source foods.^22^ Additionally, 79.5% of respondents reported being worried that they would run out of food in the past month, 13.9% reported skipping a meal in the past month and 3% reported going for an entire day without eating in the past month.^22^ Similar results were found when the cross-sectional phone survey was administered in Addis Ababa from August to September. For staples, pulses, fruits, vegetables, and animal source foods, 93.3%, 90.8 %, 59.7%, 78.7%, and 91.6 % of households reported increased food prices from the start of the pandemic, respectively.^22^

Similarly, the Ethiopian Public Health Institute and World Food Program reported that the cost of a healthy diet increased by about 17% in Amhara and 16% in Oromia between January and October 2020.^24^ A vegetable value chain survey conducted by IFPRI on value chains connecting farmers in East Shewa zone in Oromia to consumers in Addis Ababa found that price changes varied substantially by crop.^25^ The prices of tomato and onion increased by 33% and 20%, respectively, while the prices of green pepper and cabbage decreased by 13% and 12%.^25^ Transportation disruptions and border closures caused a decline in the import and the supply of some crops.^25^ For example, the import of onions from Sudan was disrupted which may have contributed to observed price increase.^25^ However, the Ethiopian dairy sector showed resilience despite the challenges of the pandemic, possibly because production systems were less dependent on market input resources compared to other systems.^26^

#### Impacts on household income, food security, and dietary consumption

Key informants indicated that many workers particularly in urban areas were unable to operate at full capacity because of the risk of transmission of COVID-19. Similarly, a phone study by IFPRI in Addis Ababa found that 64% of respondents reported a decrease in income relative to their standard income in July 2020^27^. Loss of incomes and jobs impacts consumer purchasing power, particularly to purchase nutritious foods^10^.

The impacts of COVID-19 on access to foods were most pronounced in the early months of the pandemic and decreased over time. Telephone surveys conducted by the World Bank in urban and rural households across all regions of Ethiopia found that while respondents were unable to buy certain foods due to market closures in April 2020, these disruptions subsided by May2020.^28^

A few studies examined how food security and dietary consumption changed throughout the COVID-19 pandemic. The survey conducted in Addis Ababa from August to September found that 54.2%, 19.9% and 15.3% of households reported worrying about running out of food, skipping a meal, and going for a whole day without eating in the past month, respectively.^22^ In line with these findings, an IFPRI survey in Addis Ababa found that Household Dietary Diversity Score (HDDS) significantly fell from a mean of 9.3 food groups just before the pandemic to 8.5 food groups by May and June 2020.^25^ However, despite the decrease, these dietary diversity scores in Addis were still much higher than in food-insecure areas in other areas of Ethiopia before the pandemic.^25^

### Impacts of COVID-19 on nutrition and health services

#### Impacts on provision, access, and utilization of health center services

The Ethiopian government has mainly relied on the country’s existing prevention-based primary public healthcare and health extension systems for healthcare provision. The former consists of 17,000 health posts and 3,700 health centers which employ three out of four Ethiopian healthcare professionals.^6^ The COVID-19 pandemic had some impact on the access and utilization of health services in Ethiopia that resolved over time. Key informants noted that although some found themselves with no means of transportation given COVID-19 restrictions, walking to health facilities was still possible in some communities. Most reported that fear of contracting the virus was the main driver of the disruption of access to services, especially during the first few months of the pandemic. Fear was reported as impacting both the demand and the supply side of care - patients were afraid to go to health centers and health professionals were reluctant to treat patients, largely due to the inadequate provision of personal protective equipment (PPE). A mixed-methods study conducted in woredas across four regions of Ethiopia similarly cited fear of infection and shortage of PPE as the primary drivers for decreased service utilization.^29^ Despite these initial disruptions to health services in the early stages of the pandemic, key informants and research reports highlighted health facilities’ activities had been gradually returning to normal especially after the relaxation of the lockdown measures in September 2020.^21^

Differences in the effect of the pandemic on key maternal and child health and nutrition services were observed between PHCUs in model woredas, which have successfully implemented a community health extension program and other national social mobilization strategies, and non-model woredas.^21^ PHCUs in model woredas were able to adopt better-coping strategies to resume their routine activities through outreach services.^29^ Health center staff joined the effort of HEW in the delivery of health services such as under 5 years of age malnutrition screening, under 2 years growth monitoring and deworming, along with COVID awareness through household visits.^29^ This was an effective strategy to provide care to mothers and children not coming to the health institutions due to fear of infection, and hence prevent undesirable health consequences of forgone care.

#### Impacts on antenatal and postnatal care for women and other services

COVID-19 negatively impacted the provision of key maternal nutrition services delivered at health centers, at the community level, and through schools. For services provided at health centers including maternal nutrition screening, iron, and folic acid (IFA) supplementation and antenatal care (ANC), the disruptions were more pronounced during the first 2 months of the lockdown (March and April), and gradually started improving afterward.^21^ A study by Workicho et.al (2021) indicates that ANC visits decreased by 20% throughout March and April 2020 compared to the data from the same period in 2019, as fear from acquiring the virus from primary health care centers (PHCs) affected health-seeking behaviors and led to a predilection to home deliveries.^29^

Phone surveys with Health Extension Workers (HEW) in rural areas similarly demonstrated a decrease in time spent on key maternal services as time spent on COVID-related activities increased.^21^ In May 2020 HEWs were spending 23% less time on breastfeeding counseling and antenatal care and 28% less time on postnatal care provision as compared to August 2019.^21^ While breastfeeding counseling and post-natal care had partially recovered in September 2020, ante-natal care continued to decrease.^21^

For services provided at the community level through health posts, disruptions due to the COVID-19 pandemic were most pronounced for activities that require close contact between individuals such as prenatal counselling for women, maternal deworming programs, and complimentary food preparation demonstrations^21^. These services are often provided at community level, through health posts. School antenatal nutrition services were also disrupted because of a three-month school closure. A key informant noted that UNICEF’s inability to provide IFA supplementation to adolescent girls during the closure was a key argument to ultimately re-open schools.

#### Impacts on child services and malnutrition

Two interviewees highlighted that the health system was not prepared to handle challenges in treating malnutrition cases among children at the beginning of the pandemic. Respondents noted that while there were no shortage of supplies or limitations in the availability of services to treat malnutrition, fear of contracting the virus translated into decreased demand for these services. According to an interviewee, approximately half a million children received malnutrition treatment pre-COVID, through the network of 20,000 PHC in Ethiopia^30^. Most rural facilities, that are prone to running out of supplies were mapped out by NGOs and were provided with the necessary resources to mitigate any disruption resulting from COVID-19 related movement restrictions.

In Ethiopia, most SAM cases are treated using an outpatient treatment program that uses ready-to-use therapeutic foods (RUTF). Prior to the COVID-19 pandemic, approximately 50% of RUTF for SAM treatment in Ethiopia was imported and the other half was supplied through HILINA enriched foods complex, a local producer. As key stakeholders were concerned about the potential disruption of the product supply from abroad during the pandemic, they shifted their strategy whereby HILINA started supplying all required RUTF. This strategy allowed treatment centers to continue to have sufficient resources for SAM treatment in the selected targeted districts. Despite this effort, treatment for SAM decreased by more than 50% compared to 2019 overall.^29^

However, another informant indicated that although COVID-19 posed challenges in the delivery of other child health/ nutrition services provided in the health centers, such as vitamin A supplementation, under 5 years old (U5) child deworming and screening for malnutrition increased in 2020 compared to 2019^21^. Services provided at the health care posts such as malnutrition screening, growth monitoring and deworming, all initially decreased up until May of 2020, however availability of services increased during the summer, leading to the reported increase compared to 2019^21^. This reflects the adaptive strategies used to reach out to vulnerable populations which include HEW directly going to the field and screening for malnutrition.^21^

#### Key policy responses

Key policy actions and the multitude of strategies were undertaken by the Ethiopian government and implementation partners to mitigate the impacts of COVID-19 on the food and health systems are shown in Table 3. Ethiopia’s main intervention was through its flagship food security program, the Productive Social Net Program (PSNP), to offset impacts of COVID-19 on food security for vulnerable households^4^. In addition to PSNP, both national and local governments managed to introduce smaller safety net approaches to respond to urgent needs of Ethiopians. Participation in the Productive Safety Net Program (PSNP), Ethiopia’s social protection program, mitigated the adverse impacts of the pandemic on food security.^4^ PSNP households only experienced a 2.4 percentage point increase in likelihood of becoming food insecure while the likelihood increased by 11.6 percentage points for non-PSNP households.^4^

Ethiopia launched a nationwide public information campaign and the Community Based Action and testing (ComBAT) campaign (Table 3). The latter had the twin goals of better detecting the levels of COVID-19 infection across woredas and raising awareness about prevention measures among the population.^31^ According to IFPRI’s COVID-19 Policy Response Portal, Ethiopia also increased its health budget by 46% in 2020 to deliver more equipment, deploy more personnel, and build more facilities. Despite early disruptions due to resource diversion, further disruptions were averted using HEW which continued to play a critical role in providing basic services primarily in rural areas (Table 3). In order to continue services when in-person interactions became more limited, the Ministry of Health developed a plan to deliver training modules and content on essential reproductive, maternal, newborn, child, and adolescent health and nutrition for HEWs using interactive voice response technology and mobile application.^32^

During the pandemic, to secure resources and formulate decisions in an agile manner, Ethiopia has benefitted from strong coordination between stakeholders from the public sector, civil society, and international organizations. International organizations such as IFPRI and UNICEF, responded to the pandemic by conducting data collection and analyses to understand the impacts of the pandemic and inform policy responses. Key informants interviewed in this study noted that civil society organizations have played a major role in communicating public health messages. In the health sector, respondents noted that multi-sectoral forums conducted through conference call or instant messaging applications allowed for coordination to continue despite social restrictions.

## Discussion

We conducted a qualitative study to determine the impacts of the COVID-19 pandemic on the food and health systems and evaluate the policies that were implemented to mitigate the impacts of COVID-19 in Ethiopia. We found some evidence of disruption of food system across multiple entry points including agricultural production, food availability and affordability, and dietary intake. Similarly, we found that COVID-19 related disruptions to the health systems and related nutrition services were present in the first few months. In the food system these included disruptions to the inputs supply due to travel restrictions and closed borders restricting trade, reduced in-person support by agriculture extension workers, income losses, increases in food prices, and the reduction in food security and consumption of less diverse diets. The implementation of innovative strategies and the leveraging of existing support services, such as the PSNP, reduced these impacts. The provision of nutrition services through health centers decreased in the early stages of the pandemic, primarily due to fear of contracting the virus. Impacts on health systems were mitigated over time as fear surrounding the virus subsided, PPE became available, and strategies were implemented to adapt to the disruptions, such as outreach and home services provided by the health extension workers, identification of most rural woredas and connecting them with the necessary resources, and the change to a local supplier for ready-to-use therapeutic foods.^21^

The impacts of COVID-19 on the health and food systems mirror many of the impacts felt around the world, particularly in sub-Saharan Africa (SSA). Within the food system, surveys conducted in Burkina Faso, Nigeria, and Zimbabwe found that, as in Ethiopia, the COVID-19 pandemic was associated with food price increases, reduced food availability, and lower dietary diversity.^22,33^ A study on the impact of COVID-19 on the production and food security of common bean smallholder farmers found the pandemic reduced access to seed, farm inputs, hired labor, and agricultural finance^34^. Similarly, a report on COVID-19’s impact on the agriculture sector in SSA cited reduced domestic demand, distribution disruptions, lack of access to raw materials and price increases as the most widespread impacts^35^.

The disruptions to nutrition services we report are also in line with similarly reported disruptions in SSA. A multi-country study, assessing the effects of the pandemic on healthcare services found that more than half (56%) of essential health services, including maternal and child services, were affected by the COVID-19 pandemic.^36^ These disruptions greatly varied based on the number of covid cases in the countries..^36^ A survey in Zimbabwe also found that over half of participants had trouble accessing growth monitoring services and medicinal drugs during the pandemic, which is in line with the impacts on these services in Ethiopia.^33^

To mitigate these impacts, governments across sub-Saharan Africa implemented macroeconomic and sector-specific policies that reflected aspects of Ethiopia’s response. For example, social protection programs were substantially scaled up across the region, with cash transfers increasing from 3% right before the pandemic to 11% in July 2020.^37^ Africa’s policies to mitigate COVID-19 impacts are estimated to cost about $38.5 billion, 2.4% of the GDP for African countries.^38^ The limited availability of fiscal resources to implement these policies in countries such as Ethiopia, Ghana, Mali, and Senegal, may lead to financial hardship in the future, especially as pandemics continue to emerge.^38^ African governments must create sustainable ways to prepare for such shocks and other emergencies including how to fund the policy responses.^39^ Investing in a resilient health care system in the face of crisis is crucial to mitigate not only short-term impacts but also long-term consequences resulting from potential forgone care.

The COVID-19 pandemic also underlines the fragility of Ethiopia’s social protection and healthcare systems. An inherent weakness of the systems is their reliance on external funding from organizations such as the World Bank and USAID, which means Ethiopia is exposed to shocks elsewhere. This vulnerability calls for the need for diversifying funding sources and developing local tax systems to fund social protection programs and healthcare service provisions in the long term.

There are several limitations to our study. First, considering that the COVID-19 pandemic is still ongoing, many of the impacts are yet to be seen and felt. Ongoing research and future reports will inform future practices to guide emergency responses. Second, the key informant interviews conducted for this study have informed our discussion, especially considering the familiarity of key informants in local contexts and the diversity of backgrounds they represent. Nevertheless, the perspectives shared remain as subjective opinions of a small group of informants. Third, when assessing impacts of the pandemic, we face difficulties disentangling them from the effects of other shocks experienced by Ethiopia, including conflicts in different parts of the country, macroeconomic instability due to a lack of access to foreign currencies, as well as natural disasters, such as locust infestations and flooding. As the pandemic continues, Ethiopia needs to maintain policy vigilance and invest in a long-term strategy that considers the cyclical nature of COVID-19 cases even when the number of cases subsides.

## Conclusion

Ethiopia has been able to respond to the impacts of the COVID-19 pandemic on the food and health systems by leveraging existing social protection and public health infrastructure. This pandemic has been an exercise for Ethiopia and a lesson for future pandemics, especially in terms of resource harnessing and mobilization, capacity building and public guidance. Besides being more prepared to tackle other pandemics in the future, Ethiopia’s successful response, as described by respondents, can set the right path to dealing with other infectious diseases that claim the lives of more locals such as tuberculosis. Working in agile environments, pursuing actions, assessing resources, setting emergency plans, and securing the population’s needs, especially in terms of nutrition, constitutes key actions to respond to future pandemics or outbreaks.

## Data Availability

This present study is a review of data and information that is already publically available through reports and publications. See paper bibliography for sources.

## Authors’ Note

This study investigates the ways in which the COVID-19 pandemic impacts the food system in Ethiopia including its effects on agricultural production, food prices, and consumer behaviors; assesses the impacts of COVID-19 on health systems and maternal, newborn, and child health services; and synthesizes lessons learned for Ethiopia and other countries facing the adverse effects of the COVID-19 pandemic as well as future emergencies.

## Acknowledgments

We acknowledge and thank the following key informants: Chris Andersen, Harvard T.H. Chan School of Public Health; Dr. Guush Berhane, Research Fellow at International Food Policy Research Institute; Stanley Chitekwe, Chief of Nutrition at UNICEF in Addis Ababa; Dr. Filippo Dibari, Public Health Nutritionist and Food Technologist at World Food Programme; Tefera Eshete, Agriculture Office, Disaster & Risk Management in East Hararghe Zone; Dr. Habtamu Fekadu, Senior Director of Nutrition at Save the Children; Dr. Kalle Hirvonen, Senior Research Fellow at International Food Policy Research Institute; Dr. Lisa Saldanha, Nutrition Specialist at the World Bank; Dr. Meera Shekar, Lead Health & Nutrition Specialist at the World Bank; Kyoko Shibata, Nutrition Specialist at the World Bank; Dr. Yifru Tafesse, Deputy Chief Executive Officer of the Ethiopian Agricultural Transformation Agency (ATA); Mr. Tamene Taye, Senior Nutrition Expert at the Ministry of Agriculture.

---

**Appendix II:** Overview/ Highlights of the COVID-19 response in Ethiopia^40–42^.

| Early Measures | Temporary Shock |  |  |  | Adjustment period |  | New Normal |  | Vaccines Rollout |  |
| --- | --- | --- | --- | --- | --- | --- | --- | --- | --- | --- |
|  | Measures increasing in severity amidst a controlled increase in cases- Disruption of health services and livelihood activities |  |  |  | Weekly confirmed cases increasing in May and June |  | With number of confirmed cases and deaths steadily increasing each week |  | “COVID-19 pandemic is spreading faster than ever” |  |
| February 7th 2020 | March 13th 2020 | March 16th 2020 | March 23rd 2020 | April 8th 2020 | May 2020 | June 2020 | July 2020 | October 26, 2021 | March 7th 2021 | March 13th 2020 |
| laboratory test for COVID-19 begins | - 1st covid case detected | -schools closed<br>-large gatherings discouraged<br>- flights suspended to 30 countries<br>- public transportation and movement between cities and rural areas restricted in several regions | 14-days mandatory quarantine for all travelers arriving | - Federal level state of Emergency declared<br>- land borders closed except for cargo<br>- carrying capacity of public transportation restricted (50%)<br>- Face required in public spaces | -Mitigation strategies implemented to improve the the provision and uptake of nutrition and health services<br>-ongoing travelers’ health screening at point of entries (POEs), and follow-up<br>-active case detection |  | -movement across regional states allowed<br>-humanitarian organizations operating without restrictions | -Schools in Addis Ababa reopen | - first 2.2 million doses of COVID-19 vaccine received | -COVID-19 vaccine rollout begins |

## APPENDIX II: Key Informant Interview Guide: COVID-19 disruptions of food systems and nutrition outcomes in Ethiopia: Evidence of the impacts and policy responses

Note: in these questions, we are not asking for the opinions of the key informants, but rather if they have any information or resources they can provide based on their role in their respective organization.

Questions would be included or excluded for each key informant based on their sector and organization.

### Impacts on food systems (including food security): How has COVID-19 impacted the food systems in Ethiopia?

- In what ways, if any, has COVID-19 impacted agricultural production and distribution of foods and food markets in Ethiopia?
- Are these impacts uniform across Ethiopia? Specifically what are the impacts in Addis Ababa and Kersa, if known?
- Has COVID-19 had any impacts on the food prices in Ethiopia? If so, what foods have been most impacted?
- How has COVID-19 impacted the income of individuals in various sectors and in agriculture?
- Has COVID-19 impacted the foods that are available or being purchased by consumers?
- How have individual consumers coped with the disruptions to the food system and income?
- How have the impacts differed across population groups (urban vs. rural, formal vs. informal sector workers, women-headed households etc.)?
- Are there certain groups who have been protected from the impacts of COVID-19?
- Are there currently any other non-COVID-19 disruptions to the food system (flooding, locust swarms, etc)?

### Impacts on the health systems: How has COVID-19 impacted the health systems in Ethiopia?

- How has COVID-19 impacted the health health services provided by hospitals, health centers and clinics for maternal and child health and nutrition in Ethiopia?
- Which health services are most impacted and which are least impacted by COVID-19?
- COVID 19 impact on access to ANC and PNC services, challenges and solutions?
- COVID 19 impact on malnutrition ? Did kids receive treatment? Were plumpy’nut available (F- 100; F-75)?
- What are the reasons for these disruptions (- strict lockdown vs. fear of contracting the virus vs.lack of money) and how long did they last? Were some facilities transformed to receive only COVID cases disrupting the provision of other essential health services?
- What are the consequences of these disruptions? (Short-term and long-term)

#### Policy responses to the impact of COVID-19 on food and health systems

- What policies or mitigation strategies has your organization implemented to address the impacts of COVID-19 on the food or health systems?
- Based on the evidence available, have these policies and strategies been successful at mitigating the impacts?
- Have there been any social protection programs (e.g. food aid, cash, school meals) implemented to mitigate the impact of COVID-19? Who have they targeted and who has implemented them?
- Have there been any gaps that have not been met by social protection programs

#### Further research and resources

- Are there any resources (including websites), data sources, or articles that we should be exploring to gather more information on this topic?
- Are there any key informants that we should reach out to that would be able to provide more information

